# UV-C decontamination for N95 emergency reuse: Quantitative dose validation with photochromic indicators

**DOI:** 10.1101/2020.07.17.20156497

**Authors:** Alison Su, Samantha M. Grist, Alisha Geldert, Anjali Gopal, Amy E. Herr

## Abstract

With COVID-19 N95 respirator shortages, frontline medical personnel are forced to reuse this disposable – but sophisticated – multilayer textile respirator. Widely used for decontamination of nonporous surfaces, UV-C light has germicidal efficacy on porous, non-planar N95 respirators when ≥1.0 J/cm^2^ dose is applied across all surfaces. Here, we address outstanding limitations of photochromic indicators (qualitative readout and insufficient dynamic range) and introduce a photochromic UV-C dose quantification technique for: (1) design of UV-C treatments and (2) in-process UV-C dose validation. Our methodology establishes that color-changing dosimetry can achieve the necessary accuracy (>90%), uncertainty (<10%), and UV-C specificity (>95%). Furthermore, we adapt consumer electronics for accessible quantitative readout and extend the dynamic range >10× using optical attenuators. In a measurement infeasible with radiometers, we observe striking 20× dose variation over 3D N95 facepieces. By transforming photochromic indicators into quantitative dosimeters, we illuminate critical design considerations for both photochromic indicators and UV-C decontamination.

## Introduction

Ultraviolet (UV) light in the UV-C wavelength range is one of three promising methods identified by the United States Centers for Disease Control and Prevention (CDC) for N95 respirator decontamination as a shortage mitigation strategy during the COVID-19 pandemic^1^. Building upon years of literature evidence demonstrating that specific UV-C doses inactivate viruses while preserving respirator fit and filtration^2–5^, UV-C decontamination of N95 respirators has become a rapidly expanding area of interest for both research and implementation^6^. However, effective UV-C bioburden reduction (while appearing straightforward) requires exquisite attention to detail in both treatment design and validation of treatment parameters. Challenges and intricacies of UV-C measurements can stymie study translation when UV-C dose measurements reporting viral inactivation are not robustly characterized. Innovation is urgently needed to introduce new measurement workflows that are both quantitatively robust and translatable across UV-C systems and facilities.

UV-C pathogen inactivation critically depends on two physical properties: wavelength and dose (or fluence), where dose is defined as integrated irradiance over the exposure time. Longer UV-C wavelengths (240-280 nm) inactivate pathogens like SARS-CoV-2 by damaging their genetic material (absorption peak near 260 nm)^7^ (Figure 1(a)); far UV-C also damages proteins^8^. Because UV-C decontamination relies upon pathogen interaction with electromagnetic radiation, efficacy depends on direct line-of-sight between the UV-C source and surface. Furthermore, UV-C irradiance (and integrated dose) is attenuated as light passes through multiple N95 material layers due to reflection, absorption, and scattering at each layer (Figure 1(b))^9^. Due to this attenuation, studies (almost all of which used 254 nm low-pressure mercury light) support ≥1.0 J/cm^2^ UV-C dose across the entire N95 surface for ≥3-log reduction of SARS-CoV-2 analogues on the majority of N95 models^2,4,10^: a 100-1000X higher dose than that required for non-porous surface decontamination^11^. Additionally, UV-C viral inactivation efficacy on N95s varies between models^3,4,9^, likely due to differences in respirator layer materials and 3D morphology (irradiance depends on the incident angle per Lambert’s cosine law^12^).

**Figure 1.**
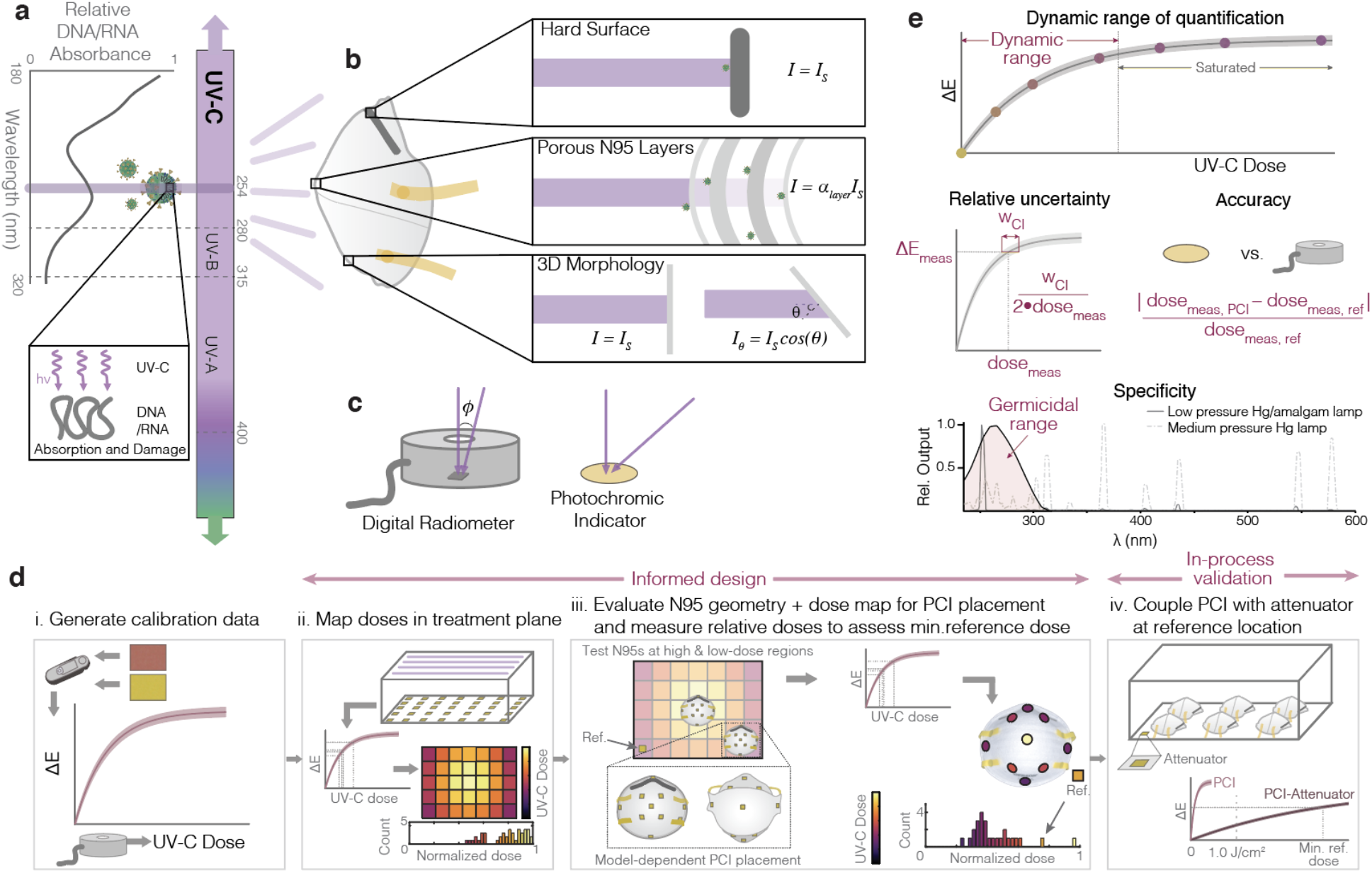
Mechanism and challenges of UV-C for N95 decontamination. (a) UV-C at 254 nm primarily inactivates pathogens by damaging genomic material (absorption peak near 260 nm). Nucleic acid absorbance spectrum modified from Voet et al.25. (b) The multilayer porous N95 materials and 3D morphology reduce the irradiance (and thus dose) available for pathogen inactivation compared to the irradiance that reaches nonporous surfaces (*I*_*S*_), like the metal nosepiece (*α*_*layer*_ represents the layer-dependent attenuation factor). (c) UV-C detectors often have angle-dependent responses that differ from the ideal cosine response expected from a surface such as flat photochromic indicators. (d) The introduced workflow allows end-users to both design and validate their UV-C systems, reducing source- and sensor-specific inaccuracies. Critically, assessment of treatment area dose nonuniformity informs N95 placement during on-N95 measurements; on-N95 measurements in turn determine minimum reference PCI doses that yield ≥1.0 J/cm^2^ to all N95 surfaces. On-N95 measurements are designed to specifically measure steep-angled or potentially-shadowed N95 regions (Supplementary Note 1). (e) Robust UV-C measurements must meet key specifications, including dynamic range of quantification (before the indicator saturates), relative measurement uncertainty (determined from error propagation from the confidence intervals on the calibration curve fit), accuracy of the measurement compared to a calibrated standard sensor, and specificity of the PCI response to the germicidal wavelength range (in order to accurately report germicidal activity). Specificity curves adapted from Kowalski^7^, Schmid *et al*.^26^, and Blickenstorfer and Aufmuth^27^. SARS-CoV-2 diagrams adapted from an image by Maya Peters Kostman for the Innovative Genomics Institute.

UV-C measurement challenges are further amplified by radiometer complexities^13^. The accuracy and relative uncertainty of digital UV-C radiometers are established through calibration to a known standard (e.g., from the National Institute of Standards and Technology)^14^; however, accuracy is dependent on sensor linearity, spectral sensitivity, and angular response^13,15^ (Figure 1(c)). Though some countries have adopted standards for inter-comparison of sensors^16^, no universal standards exist. Consequently, there is large variability between sensors in environments differing from the calibration setup, causing reproducibility challenges without meticulous detail in reporting. Furthermore, these instruments are costly, limited, low-throughput, and bulky, precluding measurements on complex 3D surfaces (which require fine spatial resolution and ideal angular response). As a result, UV-C dose is often not robustly characterized, and relative doses over the 3D N95 surface have not yet been empirically quantified.

Photochromic, color-changing UV-C indicators (PCIs) for surface decontamination are commercially available and address challenges presented by digital sensors. Due to their low cost and small, flexible form factor, PCIs are ideal for characterizing UV-C uniformity and have been applied for this characterization in hospital rooms^17^. PCIs are intended for qualitative validation; however, there has been effort to quantify color change (a topic of broader interest^18–23^) to characterize water sterilization reactors^24^.

In this work, we introduce a novel PCI-based dose quantification workflow (Figure 1(d); details in Supplementary Note 1) for informed design and validation of UV-C decontamination systems. We first demonstrate that PCI color quantification can yield UV-C-specific quantitative dose measurements with high accuracy (Figure 1(d, i)). We then use this relationship between color change and UV-C dose to show how PCIs can be implemented by end users: high throughput dose mapping within the treatment plane (Figure 1(d, ii)), combined with assessment of dose distribution across the N95 surface (Figure 1(d, iii)) allow PCIs to highlight critical locations to monitor (both on-respirator and on the treatment plane) for informed design. Relative dose measurements using PCIs can then be made on N95s positioned in the identified treatment locations (Figure 1(d, iv)) in order to establish the minimum color change reference PCIs on the treatment plane must undergo for all N95 surfaces to receive ≥1.0 J/cm^2^. Finally, we study how the addition of optical attenuator materials in front of the PCIs can extend the quantifiable UV-C dose range to ≥1.0 J/cm^2^ (Figure 1(d, iv)), enabling the final critical step of the workflow: in-process dose validation during every decontamination cycle.

## Results and Discussion

UV-C dose measurements are frequently the only link between viral inactivation studies and implementation of each decontamination cycle. Decontamination efficacy and safety consequently depend on robust UV-C measurements, defined by several critical metrics (Figure 1(e)) for which we have defined marginal and ideal values (Supplementary Table S1). Here, we introduce a new technique using PCIs to address three critical challenges hindering UV-C decontamination processes: (1) accurate and high-throughput characterization of the UV-C treatment plane (Figure 1(d, ii)), (2) spatially-resolved dose quantification across complex 3D structures placed within the treatment plane (Figure 1(d, iii)), and (3) translatable and reproducible in-process measurements to validate the dose of ≥1.0 J/cm^2^ delivered to all N95 surfaces during every UV-C treatment cycle (Figure 1(d, iv)).

### Novel photochromic indicator quantification accurately assesses spatial nonuniformities in UV-C treatment systems

PCIs have the potential to fill three urgent gaps in UV-C dose validation; however, a quantitative rather than qualitative readout strategy is required. To assess the indicators’ suitability for contributing to informed design of UV-C treatment processes, we introduce a novel quantification workflow and demonstrate the capability to capture spatial heterogeneity within a UV-C treatment system from a single exposure. We first assessed whether UV-C dose could be quantified from the color change of commercially-available PCIs; quantification relies upon distinct, reproducible color change that follows a known, predictable relationship. Measurement of color differences between the sample and a reference (rather than absolute colors) improves quantification robustness as the difference between two colors measured under the same conditions is less sensitive to many confounding effects^18,19^. To test whether two models of commercial PCIs (Intellego UVC 100 Dosimeter Dots: ‘PCI1’, and UV Process Supply UV-C Intensity Labels: ‘PCI2’) could meet the specifications of Supplementary Table S1, we exposed them to UV-C doses measured with a calibrated radiometer, quantified their color using an RM200QC spectrocolorimeter (outputting single-point L*a*b* values), and computed the CIEDE2000^28^ industry-standard color difference (ΔE) from an unexposed indicator as a function of UV-C dose (Figure 2(a)). We see comparable dose-response data for other metrics of color difference (Supplementary Note 2 and Figure S1) as well as good reproducibility between batches of PCI1 (Supplementary Figure S2). Both PCI models showed visually-discernable color change up to ∼0.15 J/cm^2^. PCI1 (Figure 2(a)) has a higher maximum ΔE of ∼45 compared to ∼25 for PCI2 (Figure 2(b)). Higher maximum ΔE will lead to lower relative uncertainty for a constant color difference measurement uncertainty.

**Figure 2.**
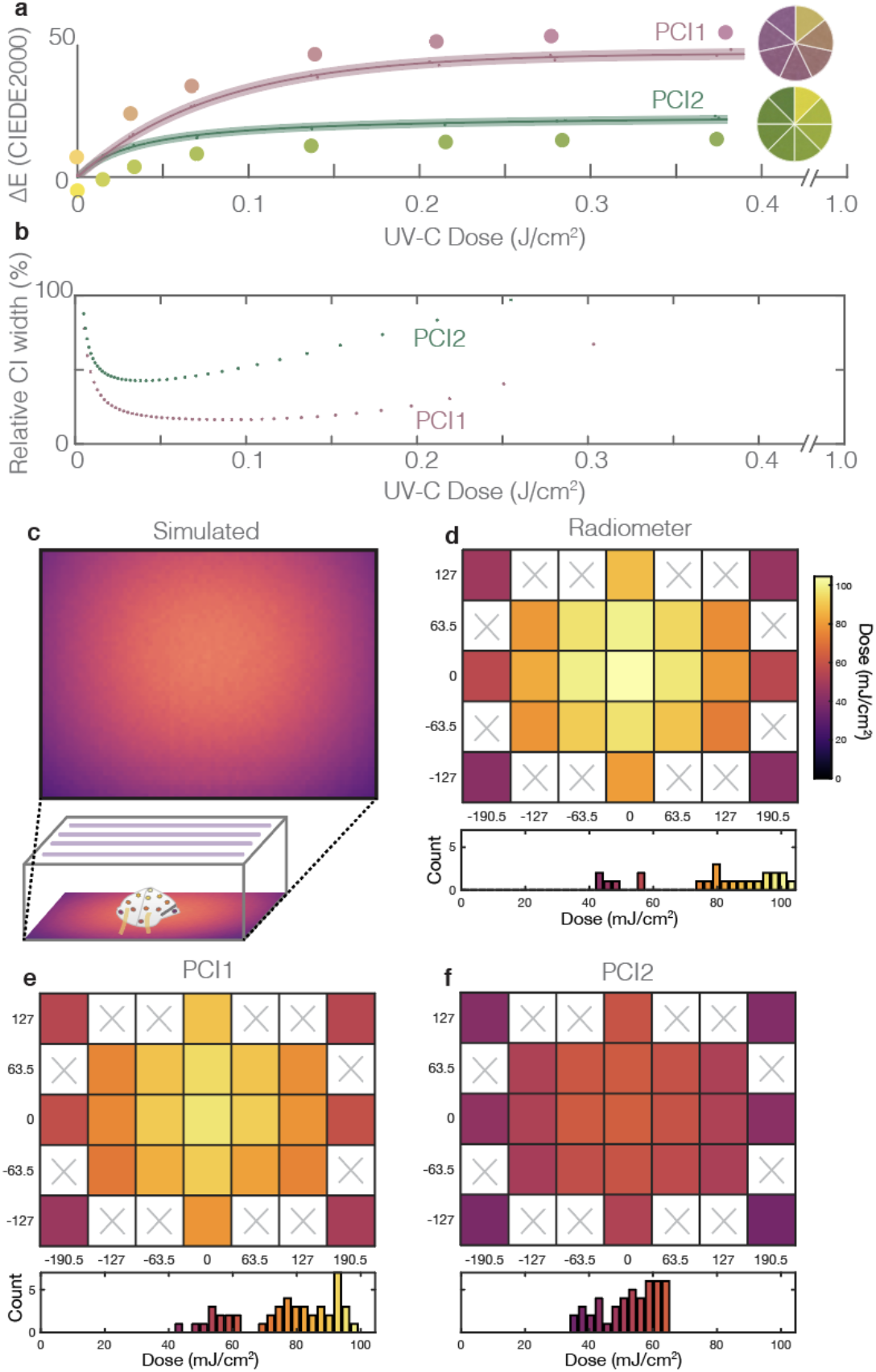
Robust color measurement facilitates UV-C dose quantification from two models of PCIs. (a) CIEDE2000 color difference between exposed and unexposed Intellego UVC Dosimeter Dot (PCI1, pink) and UV Process Supply UVC Intensity Label (PCI2, green) as a function of UV-C dose. Dose-responses for PCI1 were fit with a calibration function corresponding to first-order reaction kinetics (Eq. 2: R^2^ = 0.998; a = 47.1 (46.1, 48.1); b = 80.4 (74.6, 86.3)). Dose responses for PCI2 were fit with a calibration curve corresponding to second-order reaction kinetics (Eq. 3: R^2^ = 0.992; a = 47.7 (45.9, 49.5); b = 0.00060 (0.00049, 0.00072)). PCI color is depicted by the RM200QC-measured color values (circles) and digital SLR camera (DSLR) image swatches in the comparison wheels. Dots denote individual PCI measurements, line denotes best fit, shaded region denotes 95% confidence interval on prediction of color change from observation of UV-C dose. (b) Relative quantification uncertainties using the PCI calibration workflow. Plots depict quantified 95% confidence intervals on measurements of UV-C dose from CIEDE2000 color difference between exposed and unexposed PCIs, normalized to and as a function of UV-C dose. (c-f) heatmaps and histograms of delivered UV-C dose to locations across the treatment plane, quantified with Zemax OpticStudio simulations (c), digital radiometer (with correction factor, mean of N=3 measurements at each location) (d), PCI1 (mean of N=2 measurements at each location) (e) and PCI2 (mean of N=2 measurements at each location) (f). The PCI2 model appears to underestimate both dose and nonuniformity. Heatmaps are plotted on the same color scale (up to the radiometer maximum measured dose). White regions with ‘x’s in (d-f) were not measured.

We next scrutinized whether fitting the ΔE vs. dose data to a calibration function could predict UV-C dose from ΔE with relative dose measurement uncertainty below the 10-20% thresholds. We fit the data to calibration functions based upon first- and second-order reaction kinetics as described Supplementary Note 3. For PCI1, we used a fit function corresponding to first-order reaction kinetics (*a, b* are fit parameters):

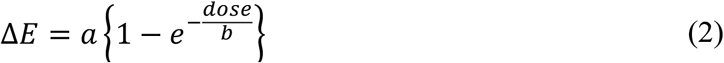

while for PCI2 we observed better goodness-of-fit with a fit function corresponding to second-order reaction kinetics (Supplementary Figure S3):

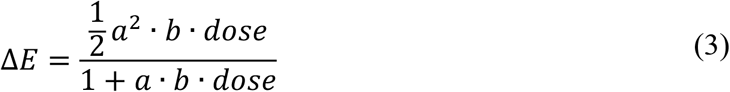

We assessed the precision of the measurement by comparing the width of the dose measurement confidence intervals (CIs) to the respective dose values (Figure 2(b)). The relative 95% CIs on UV-C doses measured with the calibration function (from known CIEDE2000 color differences) were considerably larger for PCI2 than for PCI1 suggesting that PCI1 is better-suited to robust UV-C quantification. At a dose of ∼0.1 J/cm^2^, PCI1 relative CI width (width of the CI divided by the measured dose) is 16.6% (8.3% relative uncertainty, meeting the <10% ideal relative uncertainty target). At the same dose, PCI2 relative CI width is 53.5% (26.75% relative uncertainty, above the marginally-acceptable 20% relative uncertainty target). PCI1 relative uncertainty remains <10% up to 0.15 J/cm^2^. The PCI1 relative quantification uncertainty, while higher than that established for calibration of UV-C radiometers under ideal conditions^29^, should still facilitate dose quantification as long as appropriate safety factors (i.e., a multiplier >1) are included in dose calculations to account for the propagated relative uncertainty and ensure minimum doses are met. Note that these PCIs are governed by reaction kinetics; thus, environmental factors (e.g. temperature and humidity) are expected to affect the rate of color change with dose^30^. Additionally, we have observed PCI color instability after exposure; thus, PCIs should be quantified immediately (within 10-20 minutes of exposure). Although the PCI1 quantifiable dynamic range of ≤0.15 J/cm^2^ is not sufficient for in-process validation (≥1.0 J/cm^2^ to all N95 surfaces^2–4^), it meets the ≥0.1 J/cm^2^ marginal threshold to assess relative doses for informed design of UV-C treatment systems. We also used the quantification workflow to quantify PCI specificity to UV-C wavelengths (Supplementary Note 4 and Figure S4). UV-C specificity is important because even near-monochromatic UV-C sources emit wavelengths >300 nm (Figure 1(e))^26^. When exposed to filtered (>300 nm) UV and sunlight, one PCI (PCI1) showed negligible color change (meeting the <5% specification), while ∼10% of the total color change of the other (PCI2) stemmed from >300 nm light from low-pressure amalgam illumination. These results demonstrate the importance of UV-C specificity characterization when assessing PCIs.

Having established a novel PCI quantification workflow, we next asked whether PCI measurements could scrutinize spatial dose uniformity within a UV-C treatment system as the first step towards informed design of N95 decontamination (Figure 1(d, ii)). Guiding principles of optics dictate that irradiance nonuniformities will be present in nearly any UV-C treatment system; however, the accuracy and reproducibility of UV-C measurements is hindered by a lack of standardization of critical sensor properties such as angular response, which can drastically impact readings^15,31^ (with system-dependent impact). In the absence of a calibration reference, the sensor angular response can be obtained (either through measurement^15^ or through the manufacturer) and used with optical modeling to estimate spatially-dependent system- and sensor-specific correction factors (as described in Supplementary Notes 5-6 and Figures S5-7). We first mapped UV-C dose within a Spectroline HCL-1500 UV-C source using simulation (Figure 2(c)) and 23 individual OAI 308 radiometer measurements (Figure 2(d)). We observed that the radiometer under-reports irradiance and dose due to its nonideal angular response (Supplementary Figure S8); the reported readings (Figure 2(d)) are post-correction. After correction, the irradiance measured near the corners of the treatment plane is ∼40% of that measured at the center. Simulation underestimates this nonuniformity, as validated using another radiometer with near-ideal angular response (Supplementary Figure S8).

We leveraged the nonuniform treatment plane irradiance to validate our quantification workflow by comparing PCI doses (quantified using the appropriate calibration curve depicted in Figure 2(a)) to corrected radiometer measurements (Figure 2(e-f)). The relative quantification error (|*dose*_*PCI*_ − *dose*_*radiometer*_|/*dose*_*radiometer*_) for PCI1 is 7%±5% (mean ± standard deviation of N=23 spatial measurements averaged across N=2 replicates), meeting the >90% accuracy target. In contrast, the relative quantification error for PCI2 is 30%±10%, failing to meet the marginal >80% accuracy target. While it is unclear why PCI2 performs so discordantly in this test, the good agreement between PCI1 and the radiometer suggests not only that our PCI quantification workflow can capture spatial nonuniformities in a single UV-C exposure (compared with 23 radiometer exposures), but also that color difference quantification should facilitate new classes of measurements not feasible with radiometers.

### Photochromic indicator quantification facilitates new types of measurements for informed design of UV-C treatment

PCI quantification facilitates measurements not possible with bulky radiometers (Supplementary Figure S9), such as dose mapping across complex 3D morphologies. To highlight the impact of our workflow, we mapped relative UV-C doses across the 3D morphology of a Gerson 1730 N95 respirator in three orientations (Figure 3(a)) informed by the treatment area dose mapping (Figure 2(c-e)). We exposed PCI1 indicators applied to the exterior and interior N95 surfaces to sub-saturating UV-C treatments (Figure 3(b-d)). Limited by PCI dynamic range, the exposure times were insufficient for decontamination, but measured relative dose delivered to different respirator regions (informed design, Figure 1(d,iii)). We observe that while system nonuniformities alone suggest ∼2.5X irradiance nonuniformity across the treatment plane (Figure 2(c-f)), on-N95 measurements show that nearly 20X disparity exists (Figure 3(b-d)). PCI2 dose heatmaps and dose values for both indicators are presented in Supplementary Figure S10 and Supplementary Table S2, respectively.

**Figure 3.**
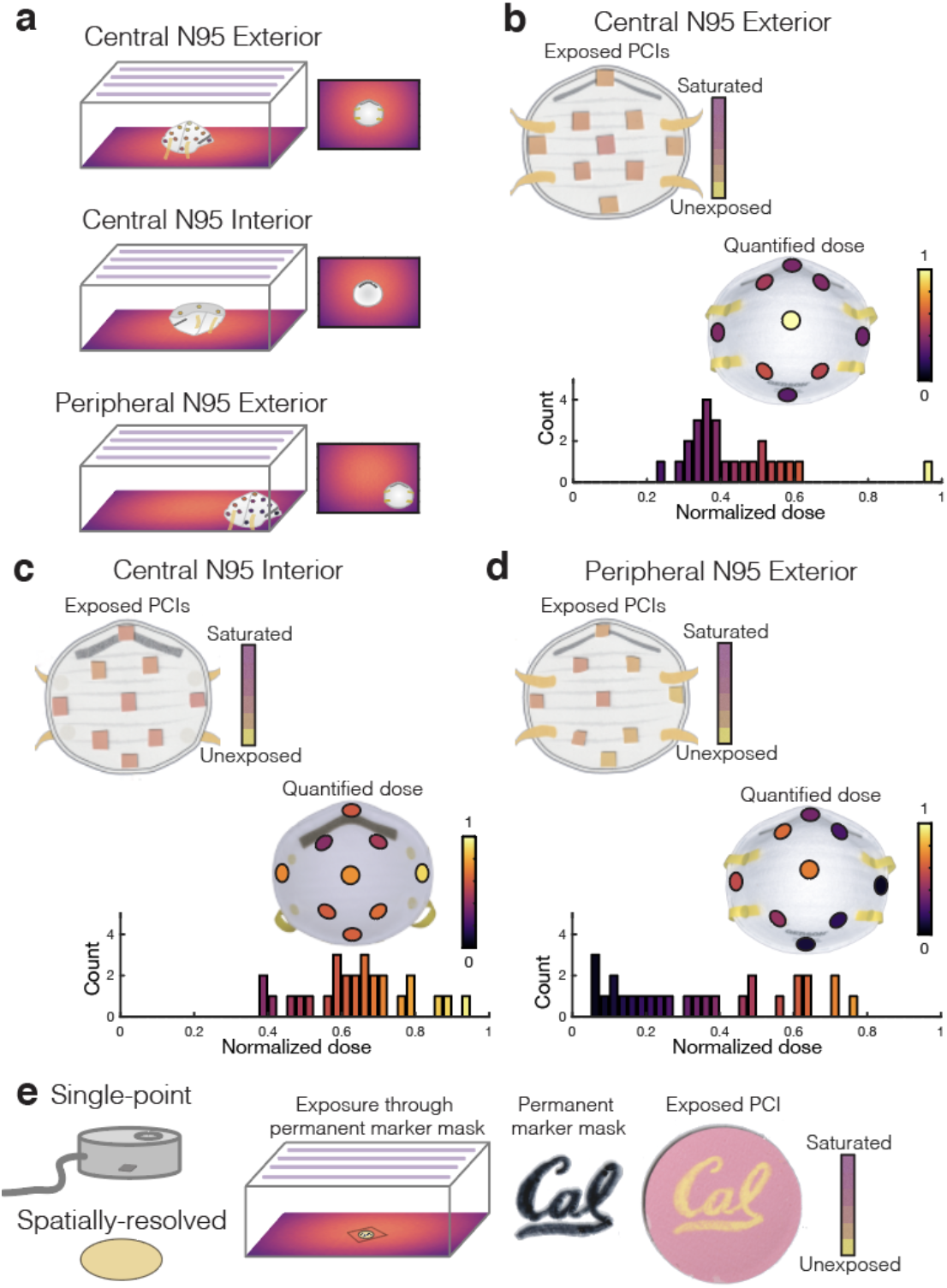
Quantifying PCIs elucidates UV-C treatment questions not measurable with radiometers. (a) Illustration of the three sets of on-N95 measurements, mapping dose across: (1) the exterior of an N95 placed in the treatment plane center (highest-dose region), (2) the interior of an N95 placed in the treatment plane center, (3) the exterior of an N95 placed in the treatment plane periphery (lowest-dose region). (b-d) Scanned images and corresponding UV-C dose quantified from PCI1 at various respirator surface locations. Although PCI color differences can appear subtle, quantification reveals substantial dose variation. Each relative dose measurement is normalized to the measurement at the apex of the central ‘convex-up’-oriented (exterior) respirator. Scanned images show a representative replicate, on-N95 heatmaps plot the mean of N=3 measurements, while the histograms below each measurement plot all individual measurements. (b) Exterior of central respirator; ∼3-4X dose difference is measured across the respirator surface. (c) Interior of central respirator. (d) Exterior of peripheral respirator. The corner-facing side of the N95 at the peripheral location is only exposed to 6.3 ± 1.3% of the dose delivered to the apex of the central respirator. (e) PCIs are 2-dimensional surface-like sensors that facilitate spatially-resolved measurements. We leveraged this characteristic to show that permanent marker (‘Cal’ pattern) on a UV-C-transparent film placed atop the PCI can mask UV-C exposure, suggesting that markings on respirators should be minimized. The PCI changes from yellow to pink as it is exposed to higher UV-C doses (see Fig. 2(a)); yellow regions correspond to areas shadowed by the marker.

The respirator morphology has a striking impact on delivered dose: even in the center of the treatment plane there are regions on the exterior (convex) N95 surface that receive only ∼25% of the dose at the apex (Figure 3(c)). There is similar but less dramatic nonuniformity present on the respirator interior (exposed concave side-up) (Figure 3(d)). Perhaps most strikingly, there are regions of a respirator in the treatment plane periphery that receive only 6% of the dose at the apex of the central N95 (Figure 3(e)). Due to the angular dependence of irradiance^12^ as well as respirator self-shadowing stemming from the 3D morphology with respect to the UV-C source, the entire N95 surface must be considered when estimating UV-C dose for decontamination; measuring the irradiance in an empty system does not sufficiently predict irradiance on the N95 surface. Scientific evidence suggests that all N95 surfaces must receive ≥1.0 J/cm^2^ UV-C dose for 3-log bioburden reduction of enveloped viruses^2–4^; however, our results show that 1.0 J/cm^2^ delivered to the apex of the central N95 in this system would result in only 0.06 J/cm^2^ applied to the side of an N95 placed in the periphery of the treatment plane. While this dose heterogeneity is certainly system- and N95 model-specific, it underscores the challenges of N95 decontamination and the critical importance of considering complex 3D geometries when designing and validating decontamination workflows.

In contrast with single-point radiometers, each PCI also records spatially-resolved doses (Figure 3(e)). As many N95 decontamination implementations track N95s using permanent marker labelling, we assessed whether such labels might shadow underlying respirator layers by positioning a pattern (‘Cal’) drawn on UV-C-transmitting film, overtop a PCI1 during exposure. We observe pattern transfer onto the indicator (Figure 3(e)), suggesting that material underneath marker labels is not as effectively decontaminated as unmarked regions. These examples of on-respirator dose quantification and spatially-resolved measurement illustrate the novel, robust measurements PCIs can provide when combined with suitable, spatially-resolved readout tools (vs. the single-measurement spectrocolorimeter), better informing UV-C treatment design.

### Device-specific calibration facilitates quantification using widely-available imaging tools

To overcome spectrocolorimeter limitations (e.g., cost, availability, and throughput) as well as capture spatial information already recorded in the PCIs, we generated and assessed device-specific calibration curves using widely-available imaging tools under controlled lighting conditions. The calibration curves were generated from raw images of PCIs acquired using a flatbed scanner (Canon LiDE-400), digital SLR camera (DSLR, Nikon D5500), and smartphone (iPhone X) (DSLR and iPhone images were acquired in a light box to provide isolation from ambient illumination). All tools captured the entire surface of both the exposed PCI as well as an unexposed reference. The resulting calibration curves were then compared to those generated with data from the RM200QC (Figure 4(a)). We observe the highest CIEDE2000 ΔE values from measurement with the cameras. Though the flatbed scanner measures the lowest ΔE values, its measurements correspond with those of the RM200QC and conveniently do not require a light box. We further assessed the squared sums of the residuals (SSE) for all fits as a measure of calibration accuracy (Figure 4(b)) and include quantification of the dose measurement relative uncertainty in Supplementary Figure S11. We observe the lowest SSE for the RM200QC, but almost all imaging approaches yield PCI1 quantification meeting the 10% ideal target relative uncertainty for a ∼0.10 J/cm^2^ dose (scanner = 10.4%). No PCI2 quantification met the marginal target, consistent with the model’s high relative uncertainty (Figure 2). The color measurement literature stresses that careful control of lighting conditions (e.g. using an enclosed light box or contact measurement) is critical in order to minimize variation induced by changes in ambient lighting^19,32^. Even under identical lighting conditions with tight control of acquisition parameters, different imaging devices have different spectral sensitivities and color processing. For this reason, device-specific calibration (using a stringent color reference chart with a range of known colors) has been proposed as an essential step for image-based color quantification for several applications^20,21,32^. A smartphone algorithm has been generated for this purpose^23^, and flatbed scanners may be a promising, accessible approach provided raw images are acquired (e.g. with third-party software). Overall, these results suggest that with rigorous characterization and proper implementation, widely-available imaging tools are appealing for spatially-resolved PCI quantification.

**Figure 4.**
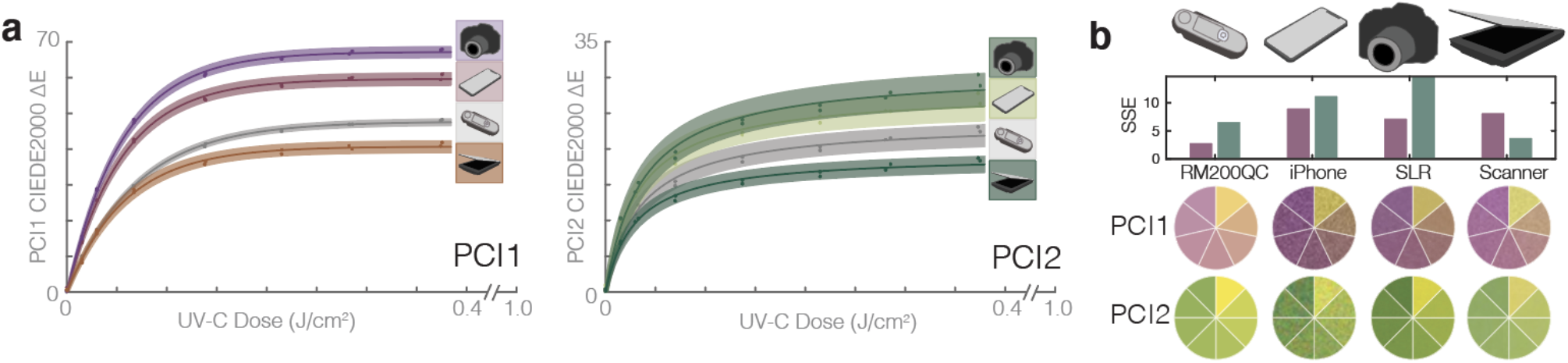
The potential applications for UV-C quantification from PCIs could be broadened using widely-available color measurement tools. (a) Comparison of calibration curve fits of CIEDE2000 ΔE from an unexposed sensor vs. UV-C dose, for PCI1 and PCI2 colors quantified using the digital SLR and iPhone X in a light box, RM200QC spectrocolorimeter, and flatbed scanner. Dots denote individual PCI measurements, line denotes best fit, shaded region denotes 95% confidence interval on prediction of color change from observation of UV-C dose. (b) Squared sum of the residuals (SSE) for each curve fit in (a), along with color comparison wheels showing indicator color at various doses as captured by the different imaging devices. Quantification of the dose-dependent relative uncertainty for each of the 8 curve fits is presented in Supplementary Figure S11.

### Optical engineering extends the quantifiable dose range for in-process validation of UV-C dose during N95 decontamination

The commercial market for UV-C PCIs has focused on hard surface UV-C decontamination processes with orders-of-magnitude lower doses than required for N95 decontamination, and thus, at this time of this study, we determined that there were no PCIs for purchase that met the dose range requirements for in-process validation of N95 decontamination. Given the benefits of PCIs over radiometers, we next assessed whether a single ‘snap-shot’ dose/irradiance measurement could be extrapolated to accurately estimate the time required for the minimum dose specification (i.e., is irradiance temporally constant?) (Figure 5(a)). Irradiance was found to substantially depend on UV-C system, warm-up status, and length of exposure (Supplementary Note 7 and Figure S12), necessitating innovation to extend the quantifiable PCI dose range for in-process validation of UV-C N95 decontamination.

**Figure 5.**
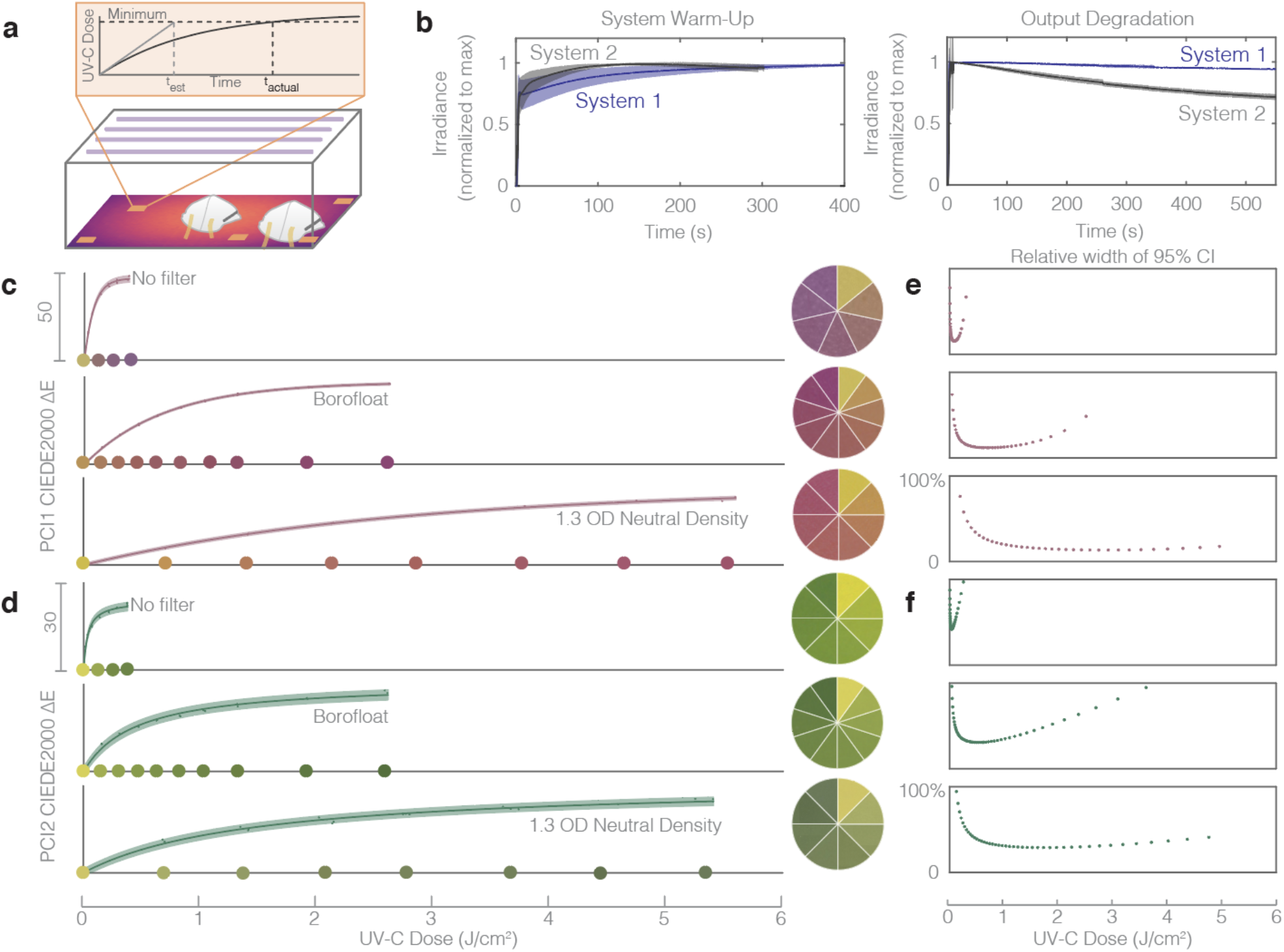
Quantitative in-process UV-C dose validation of N95 decontamination cycles using PCIs could be feasible using optical attenuation to extend the dynamic range beyond 1.0 J/cm^2^. (a) Treatment times calculated from single ‘snap-shot’ dose measurements inaccurately assume constant irradiance over time by disregarding bulb warm-up and thermal output degradation during exposure. (b) Two near-identical UV-C exposure systems have significantly different output profiles over time, both during warm-up (left), and during long exposures (right). Plots depict mean (line) and standard deviation (shaded region) of the measurements reported in Supplementary Figure S12. (c-d) 1.1 mm thick BorofloatTM glass and 1.3 OD neutral density filters extend the dynamic range of PCI1 (c) and PCI2 (d) sensors. PCI1 no filter R^2^ = 0.998, a = 47.1, b = 80.4. PCI1 with BorofloatTM filter R^2^ = 0.999, a = 47.3, b = 699. PCI1 with 1.3 OD neutral density filter R^2^ = 0.998, a = 44.2, b = 2728. PCI2 no filter R^2^ = 0.992, a = 47.7, b = 0.00060. PCI2 with BorofloatTM filter R^2^ = 0.990, a = 61.8, b = b = 3.63×10^−5^. PCI2 with 1.3 OD neutral density filter R^2^ = 0.992, a = 62.7, b = 1.10×10^−5^. Color comparison wheels show PCI colors corresponding to doses marked along the x-axis, except in “No Filter” cases due to space constraints on the x-axis. Dots denote individual PCI measurements, line denotes best fit, shaded region denotes 95% confidence interval on prediction of color change from observation of UV-C dose. The relative width of quantified 95% confidence intervals on UV-C dose measurements from CIEDE2000 color difference between exposed and unexposed samples of PCI1 (e) and PCI2 (f), as a function of UV-C dose, plotted to the right of corresponding dose-response curve.

There are two potential solutions to extend the PCI dose range: (1) engineer the indicator (e.g. modified chemistry), or (2) engineer the system surrounding the indicator. Towards (2), we assessed the capacity of several materials to attenuate the UV-C reaching the indicator to a sufficient degree to facilitate dose quantification beyond 1.0 J/cm^2^ (Figure 5(c-d)). We observe that commercial UV neutral density filters (1.3 OD) extend the quantifiable dose (<10% relative measurement uncertainty on the PCI1 measurements) from 0.15 J/cm^2^ up to 5.0 J/cm^2^ to meet the in-process validation dose specification (Supplementary Table S1). After validating that the quantifiable dose range could be extended beyond 1.0 J/cm^2^ with attenuation, we investigated a less expensive, widely-available glass option. Although standard borosilicate or soda-lime glass slides and cover slips block UV-C and are thus not appropriate filter options, 1.1 mm Borofloat™ glass offers much higher UV-C transmission^33^ and is available in a range of sizes and shapes from multiple suppliers. The Borofloat™ glass filters extended the quantifiable range of both PCIs beyond 1.0 J/cm^2^ (Figure 5(c-d), with the PCI1 measurement uncertainty remaining below 10% up to 1.5 J/cm^2^ (below 20% up to 2.0 J/cm^2^).

The dose-response curves presented in Figure 5(c-d) illustrate how precise optical attenuators can be coupled with PCIs to extend quantifiable dose range; however, the implementation of this type of approach requires careful characterization. First, before implementing any filter material, both the filter-to-filter transmittance variability and the transmittance change with UV-C exposure (e.g. due to solarization^34^) need to be characterized. Transmittance variability contributes to the uncertainty on any filtered dose measurement, while transmittance changes determine the usable lifetime of a filter (single-use vs. reuse). Second, the filter transmittance angular response needs to be assessed. Angular response can vary from the ideal cosine response based on a number of factors, including the angle-dependent optical path length through the attenuator or the angle-dependent reflectance. For a glass filter, these can be modelled using the Beer-Lambert law coupled with the Fresnel equations and Snell’s law^35^. Filter angular response could introduce similar measurement artefacts to those reported for the uncorrected radiometer in Supplementary Note 5 and thus should be understood prior to implementation. Third, filter materials should either be combined with a validated UV-C-specific photochromic indicator or have well-controlled transmission spectra to avoid a filter-indicator pair that primarily detects non-germicidal UV-A or UV-B wavelengths^7,36^. Many common materials such as glass and plastics have lower UV-C transmission than that at longer wavelengths^7^, and thus can filter out the critical wavelength range to be measured. The ideal filter would block all non-germicidal light and attenuate UV-C; however, as solarization rapidly changes transmittance and translatability requires inexpensive, widely-available materials, the ideal filter is not easily attainable. Neutral density or even higher UV-C (than longer wavelength) attenuation are acceptable when coupled with UV-C specific PCIs. Although important properties of the filter-indicator pair need to be characterized prior to implementation, the extension of quantifiable dose demonstrated in Figure 5(c-d) illustrates how Borofloat™ glass or other attenuating materials can help to address the urgent need for in-process UV-C validation, complementing future innovation in photochromic chemistry.

## Conclusions

Quantifying color from PCIs addresses urgent needs in UV-C dose measurement for (and beyond) N95 respirator decontamination. By tailoring established color measurement protocols to PCIs, we designed and validated a photochromic quantification workflow and then applied it to conduct measurements not robustly quantifiable with existing tools. Novel aspects of our workflow include quantifying CIEDE2000 for PCIs, implementing calibration informed by chemical kinetics, and quantifying PCI dose measurement uncertainty. Our workflow quantified performance specifications and revealed that while performance was highly PCI model-dependent, one indicator model met all specifications for informed design of UV-C N95 treatment systems: UV-C dose measurement range up to 0.15 J/cm^2^ with relative measurement uncertainty of 8.3% at 0.1 J/cm^2^, <5% response to UV-A/UV-B, and >90% accuracy compared to a calibrated digital radiometer. Our workflow enabled on-respirator dose quantification, identifying nearly 20X dose nonuniformity across different N95 surface regions within a treatment system. As a result, the target dose delivered to the treatment plane within the UV-C system may need to be much higher than 1.0 J/cm^2^ to ensure that all N95 surfaces are exposed to >1.0 J/cm^2^. Because these dose nonuniformities across the N95 surface are model- and configuration-dependent, each treatment system should be characterized with the N95 models *in situ* for informed design of UV-C N95 treatment processes. PCI calibration curves for widely-available imaging tools like flatbed scanners, iPhones, and DSLRs also meet minimum performance specifications and facilitate accessible, spatially-resolved dose measurements.

Finally, PCI dynamic range can be extended by coupling with optical attenuators of known transmittance. Although important open questions remain for these attenuators (such as optical transmission stability and angular response), filter-coupled PCIs are promising for high-throughput in-process dose validation for UV-C N95 decontamination. We anticipate that the PCI quantification workflow will be widely applied to meet the current urgent validation need, facilitating (1) informed design of UV-C treatment protocols to ensure that all N95 surfaces are exposed to the minimum dose, (2) in-process dose validation of each cycle, and (3) robustness characterization of new PCI materials.

## Data Availability

All reported data are available from the corresponding author upon reasonable request.

## Acknowledgments

This urgent study could not have been completed so quickly without tremendous support from a large number of generous individuals. We sincerely thank C. Cameron Miller, PhD, Maritoni Litorja, PhD, and Thomas Larason, MS, from the Optical Radiation Group at NIST, as well as Clarence Zarobila, MS, from the Remote Sensing group at NIST for developing the virtual calibration approach. We are immensely grateful to Margaret Belska, Susan Farnand, PhD, Robin Jenkin, PhD, Elaine Jin, PhD, Norman Koren, MA, Ronnier Luo, PhD, Michael Pointer, PhD, Jackson Roland, Eric Walowit, Stephen Westland. PhD, and Dietmar Wüller, Dipl.-Ing., for helpful discussions on color quantification. We appreciate contributions from Geoff McConohy, Mazzin Elsamaloty, and Josh Molho, PhD, towards preliminary attenuator characterization. We are grateful for the generosity of Intellego, UV Process Supply, and Precision Glass and Optics in providing critical materials for the study, and gratefully acknowledge helpful discussions and generous donation of time and materials from Daniel Tristan, Gustavo Garcia, Betty Cheng, MS, Philip Chau, and Mike Fleming from Spectro UV and Spectronics Corporation. We thank Ronald Moody and Evan Palmer from Optical Associates, Inc. (OAI) for support and acquiring angle-response characterization data for our purchased sensor. We would also like to thank Kyle Balston, MASc and Ellen Plane, MLA, MCP, for support and helpful discussions regarding data analysis and experimental setup. Finally, we acknowledge the other members of the N95DECON scientific consortium and medical/external partners (especially Benjamin P. Sullivan, MS, Jonathan Posner, PhD, Martin Purschke, PhD, Mark Schweitzer, and Mike Marschner) for helpful needs-finding discussions. We gratefully acknowledge funding support from the University of California, Berkeley College of Engineering Dean’s COVID-19 Emergency Research Fund, NIH training grant under award #T32GM008155 (A. Su and A. Geldert), National Science Foundation Research Fellowship Award #DGE 1106400 (A. Su), National Defense Science and Engineering Graduate (NDSEG) Fellowship (A. Geldert), the Natural Sciences and Engineering Research Council of Canada (NSERC) postdoctoral fellowship (PDF, to S.M. Grist. and postgraduate scholarship (PGS-D 487496, to A. Gopal), and the Chan Zuckerberg Biohub Investigator Program (PI: A.E. Herr).

## Author contributions

These authors contributed equally: Alison Su, Samantha M. Grist A. Su, S.M. Grist, and A.E. Herr conceived the project. S.M. Grist and A. Su designed the experiments. A. Su conducted the experiments with contributions from A. Geldert. S.M. Grist designed and implemented the quantification workflow and analyzed the data with contributions from A. Su and A. Gopal. S.M. Grist and A. Su cowrote the manuscript with input from all authors.

## Online Methods

### UV-C sensors

A Model 308 data-logging UV radiometer equipped with a 254 nm sensor (Optical Associates, Inc., OAI) was used for all irradiance measurements. An ILT1254/TD UV-C (International Light Technologies, ILT) radiometer with a near-ideal cosine angular response was used for secondary validation of irradiance measurements. Both radiometers are NIST-traceable and were calibrated within 2 months of data collection. Dose was calculated from irradiance data measured by the OAI meter and data logging software over the exposure time: 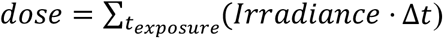.

### UV-C sources

Two different Spectroline UV treatment systems (same dimensions) were used as the UV-C source for all experiments. The XL-1500 Spectrolinker (“System 2”) was equipped with six low-pressure mercury bulbs (BLE-1T155, uvebay.com). In order to record the dose delivered in this enclosure using the radiometer, the OAI meter was wrapped in UV-C blocking material (PVC) and placed along the back wall of the chamber. This meter was plugged into a Microsoft Surface Pro tablet wrapped in multiple layers of UV-C blocking materials positioned on the left-hand side of the chamber floor. The tablet was controlled remotely using TeamViewer to record irradiance values over time. All photochromic indicator dose-response curves were measured near the center of the chamber, beside the Surface tablet. In addition, a Spectroline HCL-1500 (with the same chamber materials and dimensions as System 2, referred to as ‘System 1’) equipped with six low pressure amalgam bulbs (BLE-1T155, Spectroline) was generously donated by Spectroline with a small notch in the door to accommodate a sensor cable. With this modified instrument, data logging could be performed with the meter and tablet outside of the UV-C chamber.

### UV-C dose-response of photochromic indicators

Commercial photochromic indicators (PCIs) marketed for UV-C detection from two different companies were assessed: UVC 100 Dosimeter Dots from Intellego (‘PCI1’) and Control-Cure^®^ UV-C Intensity Labels (N010-004) from UV Process Supply (‘PCI2’). Dose measurements were quantified by integrating irradiance measurements logged by the OAI radiometer over time using a custom Python script. PCIs were placed on a plastic container of similar height to the sensor (∼5/8”). The irradiance at the PCI location was verified to be within 0.01 mW/cm^2^ of the irradiance at the radiometer location prior to measurements. PCIs were cut into pieces and a single sample was placed on either the digital sensor or plastic container and exposed during bulb warm-up to serve as a saturated reference. D65 L*a*b* measurements of both saturated and unsaturated reference PCIs were recorded using an RM200QC spectrocolorimeter (X-Rite). After bulb warm-up, sample PCIs were irradiated for a set amount of time using the “time” operating mode of the UV-C treatment system. After UV-C exposure, the color of the exposed PCI was immediately (within 5 minutes) assessed using the RM200QC spectrocolorimeter (set to report the average of three measurements of each sample).

### Quantifying dose-response curves of photochromic indicators

D65 L*a*b* measurements of PCI color assessed using the RM200QC spectrocolorimeter, along with UV-C doses (integrated from irradiance measurement logs of the radiometer readings: 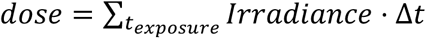 were compiled into a spreadsheet format using custom Python scripts, and then analyzed in MATLAB^®^ using scripts custom-written for this application. In order to minimize the impact of imaging/measurement conditions on the PCI color measurement, color difference from an unexposed PCI was assessed in all cases, rather than absolute PCI color. There are a range of color difference metrics^18^, and for this work we quantified and compared several.

CIELAB (1976) color difference was computed as the Euclidean distance between the L*a*b* values of the exposed (E) and reference (unexposed, R) PCIs:

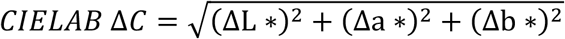

ΔL *, Δa *, and Δb * are the differences between the exposed and reference sensor L*a*b* coordinates. Similarly, the L*a*b* colors were converted to the RGB color space using built-in MATLAB functions and the Euclidean RGB color difference was computed as:

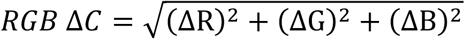

Similar as in the L*a*b* color space, ΔR, ΔG, and ΔB are the differences between the exposed and reference RGB coordinates. We also plotted, as a function of exposure dose, the differences in individual components ΔR, ΔG, and ΔB, as well as differences in lightness (ΔL *= *L* *_*E*_− *L*_*R*\*_), chroma (ΔC *= *C* *_*E*_− *C* *_*R*_), and the CIE 1976 Metric Hue Difference 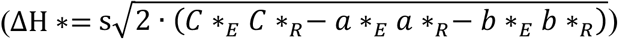, where *s* = 1 if *a* *_*R*_ *b* *_*E*_> *a* *_*E*_ *b* *_*R*_ and −1 otherwise^18^.

Finally, we computed the sets of equations for the CIEDE2000 ΔE color difference, as defined by Luo, Cui, and Rigg^28^. Our MATLAB implementation of CIEDE2000 was tested using the example color pairs presented by Luo, Cui, and Rigg^28^, and found to yield the reported ΔE values for the 10 sample-reference pairs.

### Extending the dynamic range of photochromic indicators

In order to assess the amount by which the dynamic range of the PCIs could be extended, two different filter materials were studied: a mounted 1.3 OD neutral density filter (NDUV13A, Thorlabs) and 1.10 mm thick Borofloat™ glass specified with 80/50 scratch/dig (Precision Glass & Optics). PCIs were placed on the plastic container underneath the filter material while the radiometer recorded unfiltered irradiance over time. UV-C transmission was measured using the OAI digital radiometer and calculated as the peak irradiance through the filtering material divided by the peak irradiance in the absence of filtering material.

### Characterizing variability across the treatment plane

An 11” × 17” paper grid with 2.5” markings was centered on the floor of the treatment plane. After bulb warm-up, the digital UV-C radiometer was placed at specified grid locations (Supplementary Figure S13) and peak irradiance was recorded over 15-20 seconds. The irradiance at the center of the treatment plane was verified to remain constant every 3-6 measurements to minimize variability caused by bulb output changes. The irradiances at all designated spatial locations were measured in triplicate. For PCI measurements, indicators were secured to the spatial locations on a copy of the 2.5” grid using double-sided tape. The grid was then inserted into the treatment system atop the master grid. The digital radiometer was placed in its designated location for data logging (Supplementary Figure S13). After exposure, PCIs were transferred to a consolidated layout for RM200QC analysis and measured within ∼15 minutes.

### Quantifying unknown doses using photochromic strips

In order to quantify unknown UV-C doses (e.g. across the treatment plane of the UV-C exposure system, or across the surface of an N95 respirator), color measurements from the RM200QC were read in from a spreadsheet into a custom MATLAB^®^ script. Previously-generated calibration datasets (CIEDE2000 ΔE measured with the same instrument vs. known UV-C dose, as described in “Quantifying dose-response curves of photochromic indicators” above) were read in and fitted with the calibration functions described in Supplementary Note 3. For each measurement, the L*a*b* color values for the exposed PCI and unexposed PCI reference (measured on the same day with the same instrument) were read in and the CIEDE2000 ΔE between this pair was computed as previously described. The UV-C exposure dose was predicted from the CIEDE2000 ΔE using the calibration curve. First, the inverse of the fit function was used to predict the dose from the color change. For the fit function corresponding to first-order reaction kinetics:

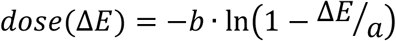

For the fit function corresponding to second-order reaction kinetics:

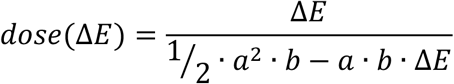

To estimate the uncertainty on the predicted dose measurement, methods for estimating uncertainties of calibrated values via propagation of error, along with uncertainties on the fitted parameters (standard deviations *s*_*a*_ and *s*_*b*_) and Δ*E* measurement (standard deviation *s*_Δ*E*_), were used to estimate the variance of the measured value 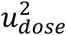^37^:

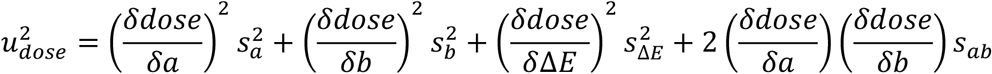

Where *s*_*ab*_ denotes the covariance between *a* and *b*. To complete this computation, *s*_*a*_, *s*_*b*_, and *s*_*ab*_ were computed from the curve fit covariance matrix, while *s*_Δ*E*_ was measured as 0.513 from the standard deviation of 20 replicate measurements of the same Pantone reference using the RM200QC. The partial derivatives of the inverse fit functions used in this computation are as described in Table 1.

**Table 1.** Fit functions, inverse fit functions, and partial derivatives used in uncertainty calculations for calibrated measurements.

| Fit type | First-order | Second-order |
| --- | --- | --- |
| Fit function | $\Delta E = a \left\{ 1 - e^{-\frac{dose}{b}} \right\}$ | $\Delta E = \frac{\frac{1}{2} a^2 \cdot b \cdot dose}{1 + a \cdot b \cdot dose}$ |
| Inverse fit function | $dose(\Delta E) = -b \cdot \ln(1 - \Delta E/a)$ | $dose(\Delta E) = \frac{\Delta E}{\frac{1}{2} \cdot a^2 \cdot b - a \cdot b \cdot \Delta E}$ |
| $\frac{\delta dose}{\delta a}$ | $\frac{-b \cdot \Delta E}{a^2 - a \cdot \Delta E}$ | $\frac{4\Delta E(\Delta E - a)}{b \cdot a^2(a - 2 \cdot \Delta E)^2}$ |
| $\frac{\delta dose}{\delta b}$ | $-\ln(1 - \Delta E/a)$ | $\frac{-2 \cdot \Delta E}{a \cdot b^2 \cdot (a - 2 \cdot \Delta E)}$ |
| $\frac{\delta dose}{\delta \Delta E}$ | $\frac{b}{a - \Delta E}$ | $\frac{2}{b \cdot (a - 2 \cdot \Delta E)^2}$ |

Ninety-five percent confidence intervals for predicted doses from each curve fit (*α* = 0.05) were predicted from the estimated variance 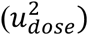 as:^38^

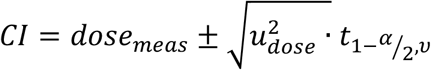

where 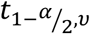 is the student’s t-inverse cumulative distribution (tinv in MATLAB^®^), and *ν* is the degrees of freedom for the calibration curve fit.

In experiments where triplicate PCI measurements of unknown doses were acquired and quantified using the calibration curve process described above, the measured doses were first equalized by correcting with a factor related to the dose logged by the radiometer during each exposure to correct for differences in the exposure time/dose between replicate measurements. To perform this correction, the doses measured from the PCI color change (as well as the confidence intervals and standard deviation of the measured value 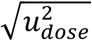) were multiplied by a target dose (constant across the replicate datasets) and divided by the logged OAI radiometer dose. After this correction for slight differences in the dose to which the PCIs were exposed, the uncertainty estimated from the standard deviation of the replicate measurements was combined with the uncertainty from the calibration fit measurements by root sum of squares:

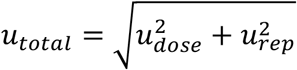

Where 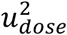 is as described above, and 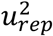 is the squared standard deviation of replicate measurements. For several datasets, dose measurement data are presented as relative doses (*dose*_*norm*_), normalized to measurements at a different location or in a different experimental setup:

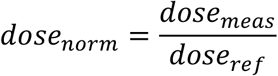

For these normalized measurements, the uncertainty is calculated from the uncertainties on both the measured and reference estimated doses via propagation of error as follows:

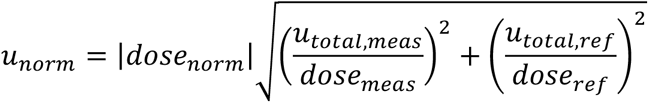

Measured doses and their uncertainties (one standard deviation, *u*_*total*_ and *u*_*norm*_) were plotted as heatmaps and histograms using the ‘inferno’ perceptually uniform, colorblind-friendly colormap, which was created by Stéfan van der Walt and Nathaniel Smith and adapted from Python’s matplotlib for use in MATLAB® by Ander Biguri^39^.

### Photochromic indicator response to non-germicidal light

A 300 nm longpass filter (#46-417, Edmund Optics) was used to assess the reactivity of the PCIs to wavelengths longer than the germicidal (200-280 nm) UV-C range. For each experiment, one PCI was placed beneath the longpass filter on top of the plastic container and one PCI was placed on the digital sensor as an unfiltered control. Post-exposure color was measured using the RM200QC. In order to assess the reactivity of the PCIs to sunlight, both models of commercial PCIs were taped to the same white background using double-sided tape and covered with black cardstock during transport outside. The exposure to sunlight began at 17:50 on May 30^th^, 2020 in Berkley, CA, USA, when the UV index was reported as 1 by Apple Weather. The color change was recorded over 5 minutes via iPhone 8 video. Both pre- and post-exposure PCIs were imaged using the Nikon D5500 and quantified using the RM200QC.

### Measuring dose received by N95 respirator surface

PCIs were affixed to the appropriate location on the surface of a NIOSH-approved Gerson 1730 N95 respirator using double-sided tape. Due to the limited dose range of the PCIs, preliminary experiments were conducted to determine an exposure time that caused all PCIs to change color within the dynamic range of the color calibration curves. For all but one condition, the exposure time was set for 8 seconds. For two exposures using PCI2 to quantify dose on a peripheral N95, the time was set for 19 seconds to take advantage of more of the PCI2 indicators’ range. These differences in exposure were compensated for in the analysis workflow described above. The respirator was positioned in its marked location within the UV-C source (either center or periphery) (Supplementary Figure 14). The straps were spread away from the respirator to minimize shadowing. For measurements of the respirator on the periphery of the treatment plane, the straps were taped together and tucked under the respirator. The OAI radiometer, with a corresponding PCI on top, was placed in its designated location for irradiance logging (Supplementary Figure S14). The color of all PCIs after exposure was recorded using the RM200QC.

### Assessing alternative imaging systems (iPhone, flatbed scanner, and digital SLR)

After each indicator exposure, the exposed indicator was imaged between unexposed and saturated references with the iPhone and Nikon D5500 within a FotodioX LED Studio-in-a-Box (FOSIAB2424, B&H) with the grey background installed. A platform was frequently inserted underneath the grey background to raise the PCIs closer to the cameras. The included diffuser sheet was cut and installed to cover the LED lights but not the top hatch. Within the Studio-in-a-Box, raw images of the PCIs were acquired using a Nikon D5500 equipped with a 40 mm macro lens or using Halide on an iPhone X at 2X optical zoom. The settings for both cameras were set manually and kept consistent within each experiment. At the conclusion of each experiment, the PCIs were scanned using VueScan (set to acquire raw images) on a flatbed scanner (LiDE 400, Canon).

### Color quantification from different imaging systems

In order to compare color quantification from the RM200QC spectrocolorimeter ‘gold standard’ tool with that from more widely available imaging devices, images of the PCIs acquired with multiple imaging devices were compared. These images were either (1) a set of images (one for each exposed PCI), each containing the exposed PCI between an unexposed and saturated PCI, with nearby white-balance region and Pantone^®^ color match to the exposed PCI (iPhone and digital SLR images), or (2) a single image of all of the exposed PCIs from a dose-response experiment, along with a single unexposed and single saturated PCI, on a white background (flatbed scanner images). iPhone and DSLR images were acquired after each exposure; scanner images were acquired once all exposures in an experiment were complete. Raw images (.DNG for iPhone X, .NEF for DSLR, .TIF for flatbed scanner) were acquired and converted to .TIF format to be read into MATLAB^®^ and analyzed using custom scripts.

In the image analysis script, each image was read in sequentially and the user prompted to draw rectangular areas over (a) the exposed PCI, (b) the Pantone^®^ match to the PCI, (c) the white region proximal to the PCI(s), (d) the unexposed PCI, and (e) the saturated PCI. In all cases, care was taken to draw a region encompassing only the region of interest (i.e., not edges, dust, or shadowed regions). For the first type of data (1: an image for each PCI), all 5 regions were denoted on each image (for each exposed PCI). For the second type of data (2: a single image for all PCIs), a single region was denoted for the white, unexposed, and saturated regions for all exposed PCIs, with only the PCI region denoted for each exposure dose (Pantone^®^ matches were not imaged in the second type of imaging workflow). After all regions on each image were selected, the average RGB value for the white region was used to white-balance and exposure-correct the image before computing the average RGB values for the other region types. The RGB value for each region was then converted to the L*a*b* color space using MATLAB^®^’s built-in rgb2lab function. RGB and L*a*b* values from the processed images were then subjected to the same processing for color difference calculations as described above for the measured RM200QC L*a*b* values in “Quantifying dose-response curves of photochromic indicators”.

CIEDE2000 color differences from an unexposed PCI, computed from each image type as well as the RM200QC measurements of the same set of PCIs, were fitted to the appropriate calibration function and plotted (along with 95% prediction intervals) as a function of exposure dose, in order to compare the relative dose-responses and calibration uncertainties measured with each tool. The squared sum of the residuals from the curve fit (SSE) for each dataset was computed and compared as a metric of calibration robustness for each color readout method.

### Visualizing reduced UV-C transmission through permanent marker ink

To demonstrate spatially-resolved measurement, we visualized the UV-C shadowing abilities of permanent marker ink. The “Cal” university logo was drawn with Sharpie® permanent marker on a UV-C-transmissive (∼82% transmittance) plastic plate sealer adhesive film. The plastic film with permanent marker logo was then placed atop a PCI1 indicator within the UV-C treatment plane and exposed to UV-C for ∼10s (applied UV-C not precisely controlled as this was a qualitative test). After exposure, the film and exposed PCI were imaged using the flatbed scanner.

### Assessing temporal fluctuations in irradiance

Irradiances over time logged using the OAI radiometer either during system warm-up or during long-exposures after warm-up were parsed from the output .txt files using a custom Python script and read into MATLAB^®^. Warm-up datasets approximated the variance that would be present in applied conditions because the time since previous use was not controlled (the datasets began with the lamps in varying states of warm-cool). Each dataset was analyzed to automatically detect the iteration (*iend*) at which lamp shutoff occurred (from the change from the previous measurement). The irradiance data were plotted until 2 measurements prior to that measurement iteration (*iend-2*). For the system warm-up datasets, warm-up rise time was computed as the time for the irradiance to rise from 10% of the maximum recorded value to 90% of the maximum recorded value. For the long exposure datasets, the output degradation was assessed by extracting the irradiance degradation slope from linear least-squares curve fitting.

The estimated time to reach 1.0 J/cm^2^ from each exposure was computed as the target dose (1.0 J/cm^2^) divided by the mean irradiance. To assess the effects of temporal instability of the lamp output, this calculation was computed for both a mean irradiance at the beginning of each exposure (taking the mean irradiance over the 10^th^ to 20^th^ iterations of the data logger) and a mean irradiance at the end of each exposure (taking the mean irradiance over the last 11 iterations prior to the detected end point (*iend*, automatically detected from lamp shutoff as described above).

## References

1. CDC. Decontamination and Reuse of Filtering Facepiece Respirators. Centers for Disease Control and Prevention https://www.cdc.gov/coronavirus/2019-ncov/hcp/ppe-strategy/decontamination-reuse-respirators.html (2020).

2. Lore, M. B., Heimbuch, B. K., Brown, T. L., Wander, J. D. & Hinrichs, S. H. Effectiveness of Three Decontamination Treatments against Influenza Virus Applied to Filtering Facepiece Respirators. Ann. Occup. Hyg. 56, 92–101 (2011).

3. Mills, D., Harnish, D. A., Lawrence, C., Sandoval-Powers, M. & Heimbuch, B. K. Ultraviolet germicidal irradiation of influenza-contaminated N95 filtering facepiece respirators. Am. J. Infect. Control 46, e49–e55 (2018).

4. Brian Heimbuch, Del Harnish. Research to Mitigate a Shortage of Respiratory Protection Devices During Public Health Emergencies. https://www.ara.com/sites/default/files/MitigateShortageofRespiratoryProtectionDevices_2.pdf (2019).

5. Lindsley, W. G. et al. Effects of Ultraviolet Germicidal Irradiation (UVGI) on N95 Respirator Filtration Performance and Structural Integrity. J. Occup. Environ. Hyg. 12, 509–517 (2015).

6. John J. Lowe et al. N95 Filtering Facemask Respirator Ultraviolet Germicidal Irradiation (UVGI) Process for Decontamination and Reuse. https://www.nebraskamed.com/sites/default/files/documents/covid-19/n-95-decon-process.pdf (2020).

7. Kowalski, W. Ultraviolet Germicidal Irradiation Handbook: UVGI for Air and Surface Disinfection. (Springer Berlin Heidelberg, 2009). doi:10.1007/978-3-642-01999-9.

8. Beck, S. E., Hull, N. M., Poepping, C. & Linden, K. G. Wavelength-Dependent Damage to Adenoviral Proteins Across the Germicidal UV Spectrum. Environ. Sci. Technol. 52, 223–229 (2018).

9. Fisher, E. M. & Shaffer, R. E. A method to determine the available UV-C dose for the decontamination of filtering facepiece respirators: UV-C decontamination of respirators. J. Appl. Microbiol. 110, 287–295 (2011).

10. Heimbuch, B. K. et al. A pandemic influenza preparedness study: Use of energetic methods to decontaminate filtering facepiece respirators contaminated with H1N1 aerosols and droplets. Am. J. Infect. Control 39, e1–e9 (2011).

11. Tseng, C.-C. & Li, C.-S. Inactivation of Viruses on Surfaces by Ultraviolet Germicidal Irradiation. J. Occup. Environ. Hyg. 4, 400–405 (2007).

12. Reifsnyder, W. E. Radiation geometry in the measurement and interpretation of radiation balance. Agric. Meteorol. 4, 255–265 (1967).

13. Bolton, J. R. & Linden, K. G. Standardization of Methods for Fluence (UV Dose) Determination in Bench-Scale UV Experiments. J. Environ. Eng. 129, 209–215 (2003).

14. Lawal, O. et al. Method for the Measurement of the Output of Monochromatic (254 nm) Low-Pressure UV Lamps. IUVA News 19, 9–16.

15. Reed, N. G., Wengraitis, S. & Sliney, D. H. Intercomparison of Instruments Used for Safety and Performance Measurements of Ultraviolet Germicidal Irradiation Lamps. J. Occup. Environ. Hyg. 6, 289–297 (2009).

16. Schmalwieser, A. W. Fifteen years of experience with standardized reference radiometers for controlling low-pressure UV disinfection plants for drinking water. Water Supply 17, 975–984 (2017).

17. Lindblad, M., Tano, E., Lindahl, C. & Huss, F. Ultraviolet-C decontamination of a hospital room: Amount of UV light needed. Burns 46, 842–849 (2020).

18. ASTM Standard D2244-16. Standard Practice for Calculation of Color Tolerances and Color Differences from Instrumentally Measured Color Coordinates. ASTM Int. (2016) doi:10.1520/D2244-16.

19. Cui, G., Luo, M. R., Rhodes, P. A., Rigg, B. & Dakin, J. Grading textile fastness. Part 1; Using a digital camera system. Color. Technol. 119, 212–218 (2003).

20. Carpenter, K. & Farnand, S. Assessing the use of smartphones to determine crop ripeness. Electron. Imaging (2020) doi:10.2352/ISSN.2470-1173.2020.12.FAIS-173.

21. Soda, Y. & Bakker, E. Quantification of Colorimetric Data for Paper-Based Analytical Devices. ACS Sens. 4, 3093–3101 (2019).

22. Singh, G., Raj, P., Singh, H. & Singh, N. Colorimetric detection and ratiometric quantification of mercury(II) using azophenol dye: ‘dip & read’ based handheld prototype device development. J. Mater. Chem. C 6, 12728–12738 (2018).

23. Yetisen, A. K., Martinez-Hurtado, J. L., Garcia-Melendrez, A., da Cruz Vasconcellos, F. & Lowe, C. R. A smartphone algorithm with inter-phone repeatability for the analysis of colorimetric tests. Sens. Actuators B Chem. 196, 156–160 (2014).

24. Solari, F., Girolimetti, G., Montanari, R. & Vignali, G. A New Method for the Validation of Ultraviolet Reactors by Means of Photochromic Materials. Food Bioprocess Technol. 8, 2192–2211 (2015).

25. Voet, D., Gratzer, W. B., Cox, R. A. & Doty, P. Absorption spectra of nucleotides, polynucleotides, and nucleic acids in the far ultraviolet. Biopolymers 1, 193–208 (1963).

26. Schmid, J., Hoenes, K., Rath, M., Vatter, P. & Hessling, M. UV-C inactivation of Legionella rubrilucens. GMS Hyg. Infect. Control 12, Doc06–Doc06 (2017).

27. Blickenstorfer, B. & Aufmuth, W. Energy-efficient crosslinking with low-pressure lamps. Adhes. Adhes. 10, 26–31 (2013).

28. Luo, M. R., Cui, G. & Rigg, B. The development of the CIE 2000 colour-difference formula: CIEDE2000. Color Res. Appl. 26, 340–350 (2001).

29. Larason, T. & Ohno, Y. Calibration and characterization of UV sensors for water disinfection. Metrologia 43, S151–S156 (2006).

30. El Seoud, O. A., Baader, W. J. & Bastos, E. L. Practical Chemical Kinetics in Solution. in Encyclopedia of Physical Organic Chemistry, 5 Volume Set (ed. Wang, Z.) 1–68 (John Wiley & Sons, Inc., 2016). doi:10.1002/9781118468586.epoc1012.

31. Larason, T. C. & Cromer, C. L. Sources of error in UV radiation measurements. J. Res. Natl. Inst. Stand. Technol. 106, 649 (2001).

32. Nixon, M., Outlaw, F., MacDonald, L. W. & Leung, T. S. The importance of a device specific calibration for smartphone colorimetry. Color Imaging Conf. 2019, 49–54 (2019).

33. SCHOTT Technical Glass Solutions GmbH. BOROFLOAT® 33 – Optical Properties. https://www.schott.com/d/borofloat/bde16ad3-70e5-48a0-b8ac-9146fcd34511/1.0/borofloat33_opt_en_web.pdf.

34. Natura, U., Ehrt, D., Naumann, K. & Glas, S. Formation of radiation defects in high-purity silicate glasses in dependence on dopants and UV radiation sources. Glas. Ber Glass Sci. Technol. 74, 23–31 (2001).

35. Furler, R. A. Angular Dependence of Optical Properties of Homogeneous Glasses. ASHRAE Trans. 98, 1–9 (1991).

36. Lytle, C. D. & Sagripanti, J.-L. Predicted Inactivation of Viruses of Relevance to Biodefense by Solar Radiation. J. Virol. 79, 14244–14252 (2005).

37. NIST/SEMATECH. 2.3.6.7.1. Uncertainty for quadratic calibration using propagation of error. NIST/SEMATECH e-Handbook of Statistical Methods https://www.itl.nist.gov/div898/handbook/mpc/section3/mpc3671.htm DOI: 10.18434/M32189 (2020).

38. NIST/SEMATECH. 4.5.2.1. Single-Use Calibration Intervals. NIST/SEMATECH e-Handbook of Statistical Methods https://www.itl.nist.gov/div898/handbook/pmd/section5/pmd521.htm#:~:text=Although%20calibration%20confidence%20intervals%20have,for%20the%20true%20average%20response. DOI: 10.18434/M32189 (2020).

39. Biguri, A. Perceptually uniform colormaps. MATLAB Central File Exchange https://www.mathworks.com/matlabcentral/fileexchange/51986-perceptually-uniform-colormaps (2020).

